# Causes of death in mental health service users during the first wave of the COVID-19 pandemic: South London and Maudsley data from March to June 2020, compared with 2015-2019

**DOI:** 10.1101/2020.10.25.20219071

**Authors:** Robert Stewart, Amelia Jewell, Matthew Broadbent, Ioannis Bakolis, Jayati Das-Munshi

**Affiliations:** King’s College London (Institute of Psychiatry, Psychology and Neuroscience), London, UK; South London and Maudsley NHS Foundation Trust, London, UK

## Abstract

The COVID-19 pandemic is likely to have had a particularly high impact on the health and wellbeing of people with pre-existing mental disorders. This may include higher than expected mortality rates due to severe infections themselves, due to other comorbidities, or through increased suicide rates during lockdown. However, there has been very little published information to date on causes of death in mental health service users. Taking advantage of a large mental healthcare database linked to death registrations, we describe numbers of deaths within specific underlying-cause-of-death groups for the period from 1^st^ March to 30^th^ June in 2020 and compare these with the same four-month periods in 2015-2019. In past and current service users, there were 2561 deaths in March-June 2020, compared to an average of 1452 for the same months in 2015-19: an excess of 1109. The 708 deaths with COVID-19 as the underlying cause in 2020 accounted for 63.8% of that excess. The remaining excess was accounted for by unnatural/unexplained deaths and by deaths recorded as due to neurodegenerative conditions, with no excess in those attributed to cancer, circulatory disorders, digestive disorders, respiratory disorders, or other disease codes. Of 295 unexplained deaths in 2020 with missing data on cause, 162 (54.9%) were awaiting a formal death notice (i.e. the group that included deaths awaiting a coroner’s inquest) – an excess of 129 compared to the average of previous years, accounting for 11.6% of the excess in total deaths.

## Background

The COVID-19 pandemic is likely to have had a profound impact on people using mental healthcare because of the heightened vulnerability of its patient populations (e.g. through cardiovascular and respiratory disorders), already-reduced life-expectancies (1), and frequently described problems accessing healthcare (2, 3). There is therefore a pressing need for information in the public domain (4, 5). This manuscript is one of a series of open-access reports, covering different elements of mental health service activity and outcomes from a large mental healthcare provider in south London, UK. The reports take advantage of relatively rapid access to source data, as well as a growing network of linked data, and will be kept updated for as long as a single site’s experiences are deemed useful.

We have previously described a 2.4-fold excess mortality in past and present mental healthcare services users during the period of the pandemic experienced in London from 16^th^ March to 15^th^ May 2020 compared to mortality for the same 2-month period in 2019 (6), as well as ethnic group differences in mortality (7). Drawing on recently linked data on cause of death for deaths up to the end of June 2020, we sought to describe numbers and distributions of recorded causes for deaths in past and current mental health service users (during the March-June period (i.e. covering the first wave of the COVID-19 pandemic experienced in London, UK) and to compare these with data for the same months from 2015-2019.

## Methods

The Biomedical Research Centre (BRC) Case Register at the South London and Maudsley NHS Foundation Trust (SLaM) has been described previously (8, 9). SLaM serves a geographic catchment of four south London boroughs (Croydon, Lambeth, Lewisham, Southwark) with a population of around 1.2 million residents and has used a fully electronic health record (EHR) across all its services since 2006. SLaM’s BRC Case Register was set up in 2008, providing researcher access to de-identified data from SLaM’s EHR via the Clinical Record Interactive Search (CRIS) platform and within a robust security model and governance framework (10). Of relevance to the work presented here, CRIS is updated from SLaM’s EHR every 24 hours and thus provides relatively ‘real-time’ data; currently CRIS contains data on over 500,000 past and current SLaM service users.

Mortality in the complete EHR (i.e. all SLaM patients with records, past or present) is ascertained weekly through automated checks of National Health Service (NHS) numbers (a unique identifier used in all UK health services) against a national spine; in addition, a data linkage has been set up and is updated annually between CRIS and death certifications held by the Office for National Statistics (ONS) which was recently updated for deaths up to the end of June. CRIS has supported over 200 peer reviewed publications to date. CRIS has received approval as a data source for secondary analyses (Oxford Research Ethics Committee C, reference 18/SC/0372).

Mortality data were extracted on all individuals contained in CRIS (i.e. past and current SLaM service users) for the period from 1^st^ March to 30^th^ June 2020, as this coincided with the first wave of the COVID-19 pandemic in London. Comparison data were extracted in an identical way for the same four months in each of 2015-2019. This extraction drew on both available sources of data on deaths, as not all recorded deaths receive certifications and there may also be delays in certification data recorded on the ONS linkage files. The following categories were applied according to the listed underlying cause of death (coded according to International Classification of Diseases 10^th^ Edition, ICD-10):

1. COVID-19 (ICD-10 U071 or U072)
2. Cancer (ICD-10 C)
3. Circulatory disorders (ICD-10 I)
4. Digestive system disorders (ICD-10 K)
5. Mental disorders (ICD-10 F)
6. Neurological disorders (ICD-10 G)
7. Respiratory disorders (ICD-10 J)
8. Unnatural/unexplained causes: these included causes of death under ICD-10 R, U, W, X and Y codes (apart from U071 and U072), as well as deaths ascertained via the NHS spine link without a cause of death listed on the ONS linkage.
9. All other causes (labelled as ‘all other disorders’ in graphical displays)

Having described total numbers of deaths in the above categories for each of the years, further descriptions were provided according to the following characteristics, defined on the 1^st^ March for each respective year: i) age group (<70y or 70+); ii) sex; iii) whether ‘active’ to SLaM (i.e. having an accepted referral or IAPT psychotherapy treatment episode) during the March-June period in question; iv) ethnic group (conflated because of numbers to White British, Black African/Caribbean, Other). Further analyses were carried out to provide more detailed breakdowns of deaths recorded as due to mental/neurological disorders, and those that were ascertained via the NHS spine link but not on the ONS linkage.

## Results

Numbers of deaths by underlying cause and year for all SLaM patients are displayed in Figure 1. There were 2561 deaths in March-June 2020, compared to an average of 1452 for the same months in 2015-19: an excess of 1109. There were a total 708 deaths in 2020 with COVID-19 as the identified underlying cause, accounting for 63.8% of that excess. The remaining excess deaths were in the unnatural/unexplained category and in the mental/neurological disorders categories, with other causes showing no consistent excess.

**Figure 1:**
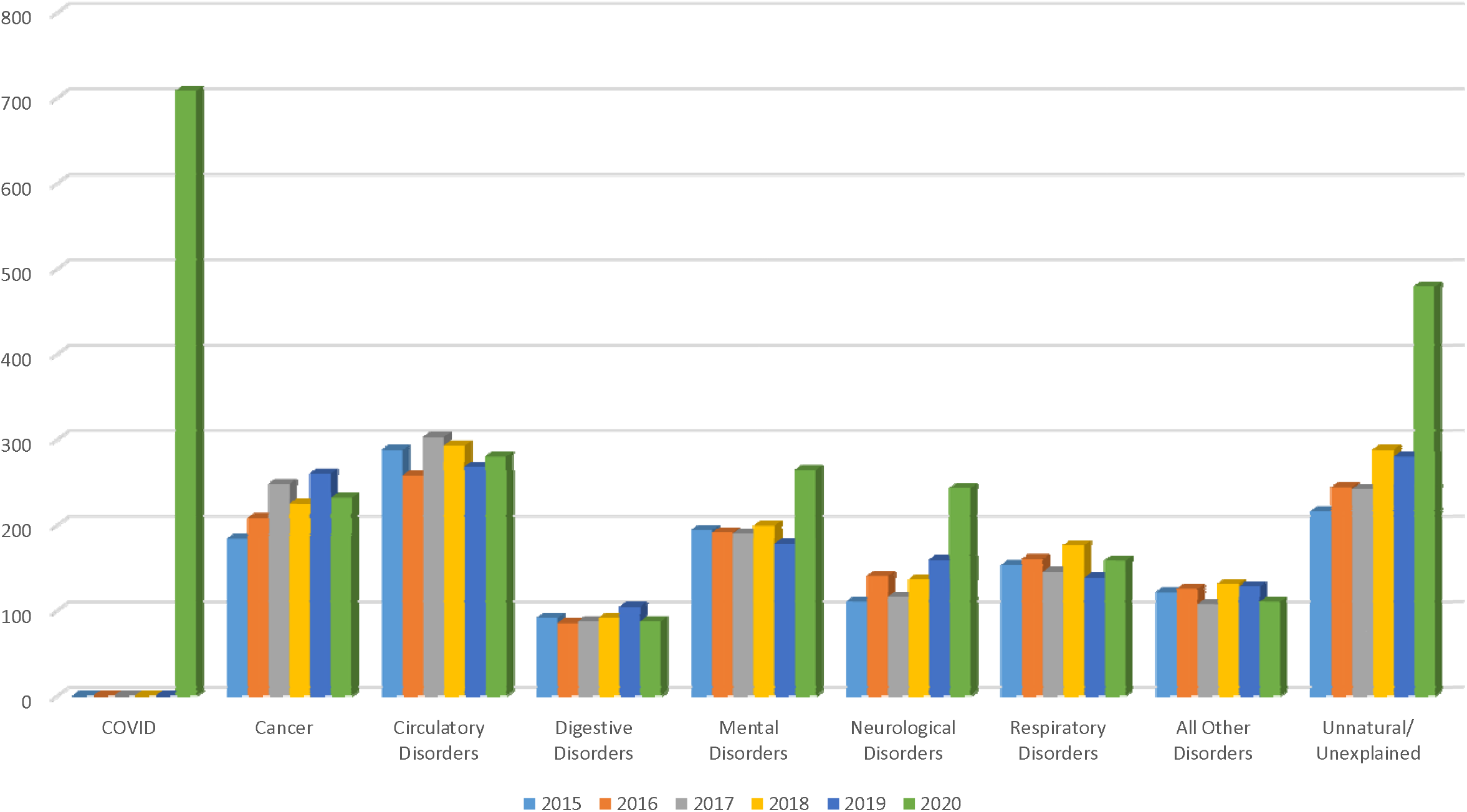
Numbers of deaths during March-June by underlying cause of death and year for all SLaM service users

On stratification by age (Figures 2a and 2b), 535 (75.2%) of deaths due to COVID-19 occurred in those aged 70+ years and the majority of deaths (and excess deaths in 2020) due to mental/neurological disorders occurred in this age range. Numbers of unnatural/unexplained deaths were similar in those aged <70 and 70+ (258 and 224 respectively), but the excess compared to previous years was more pronounced in the older age group.

**Figure 2a:**
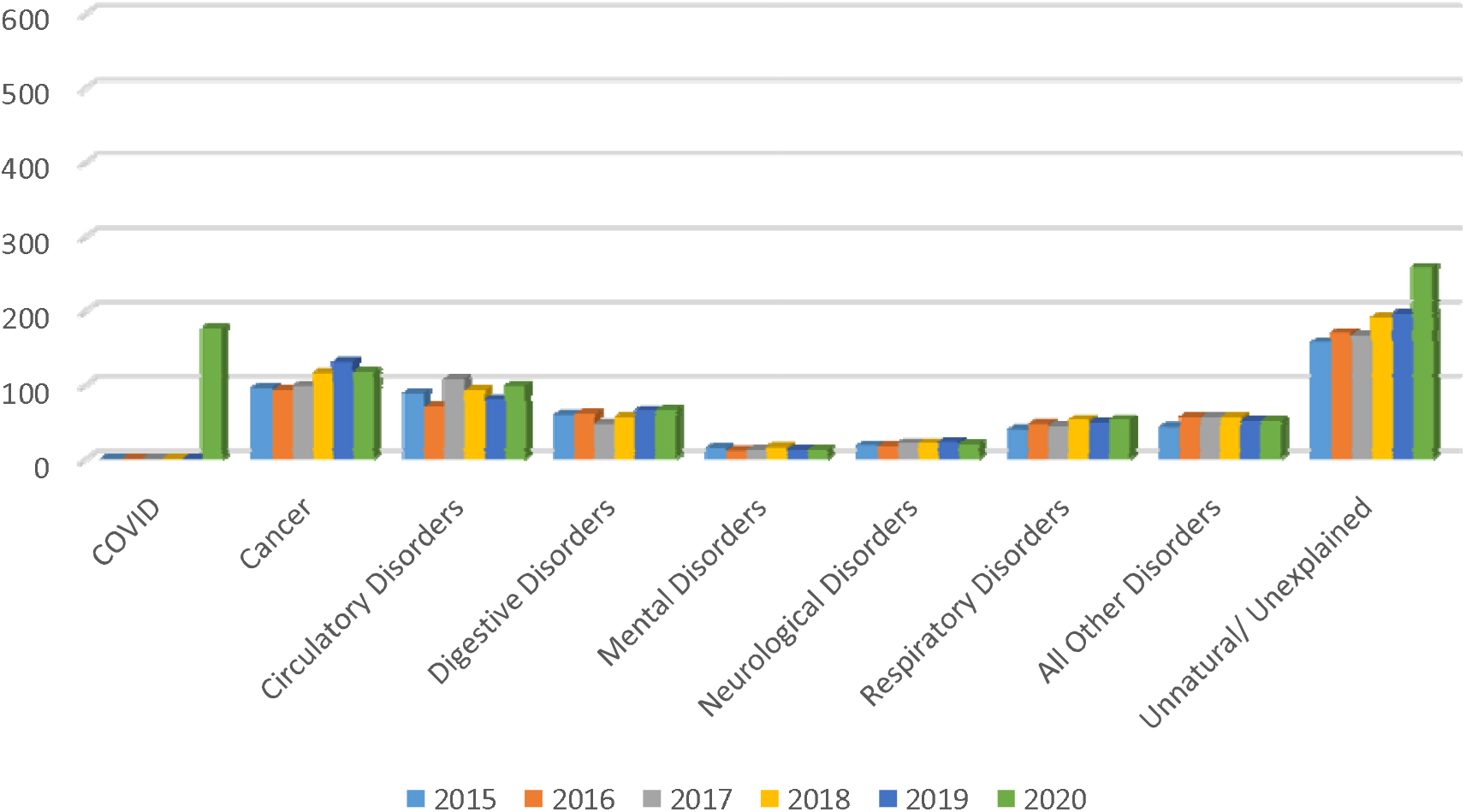
Numbers of deaths in SLaM service users aged <70 from March to June by underlying cause of death and year

**Figure 2b:**
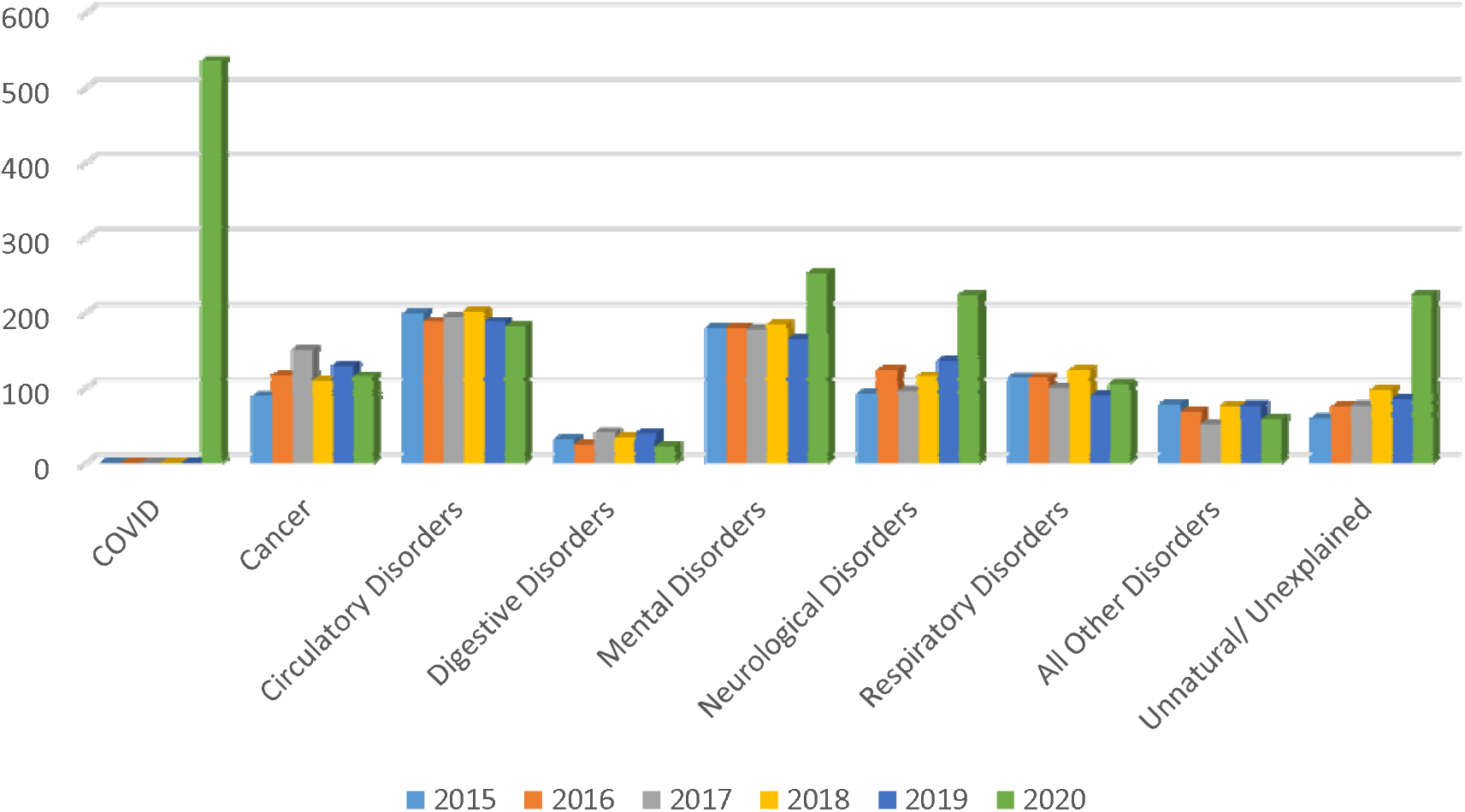
Numbers of deaths in SLaM service users aged 70+ from March to June by underlying cause of death and year

On stratification by sex (Figures 3a and 3b), there was a male excess in deaths due to COVID-19 and in unnatural/unexplained deaths, although 2020 differences compared to previous years were present for both groups. Excess deaths due to mental/neurological conditions were more marked in females than males.

**Figure 3a:**
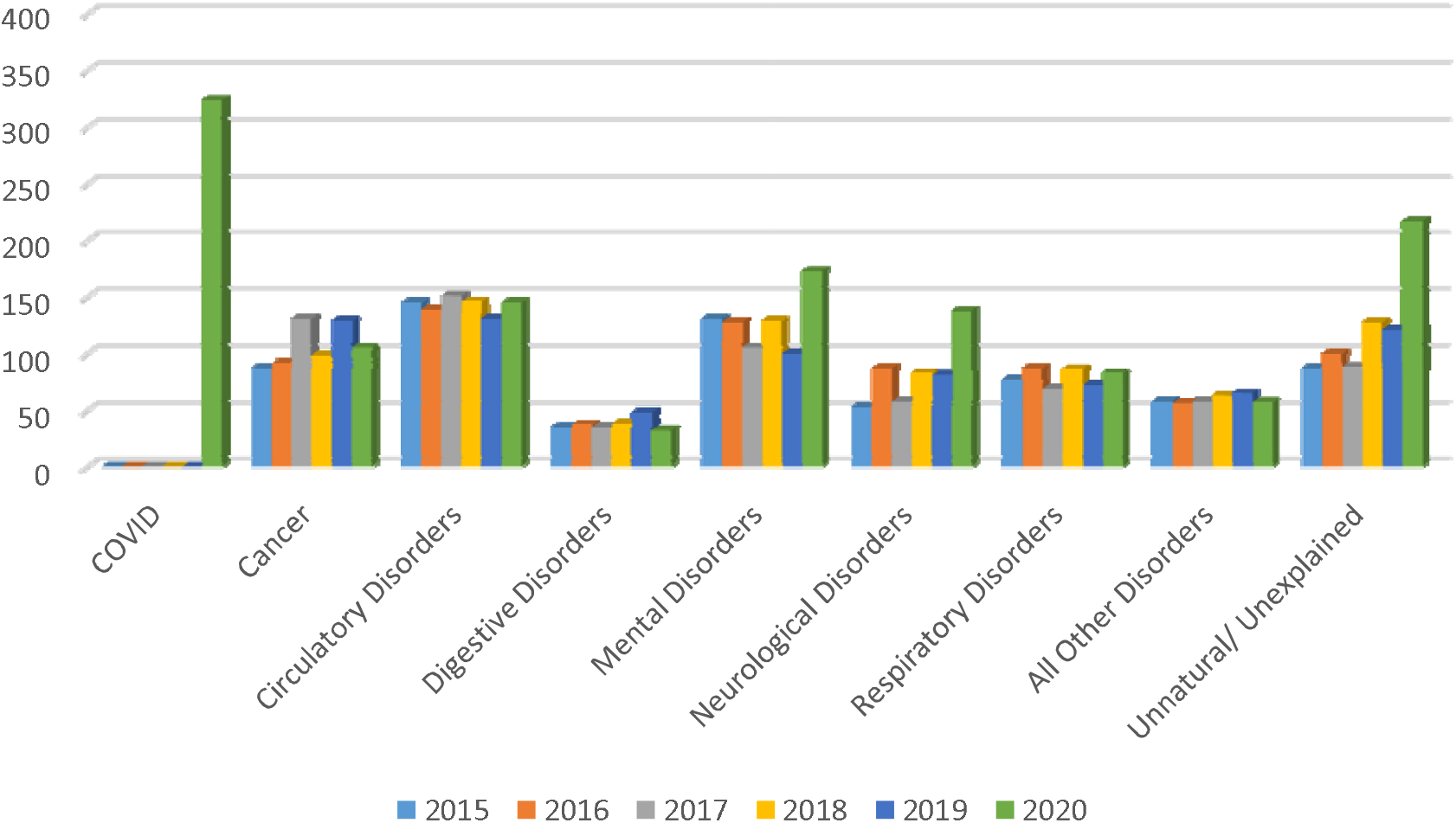
Numbers of deaths in female SLaM service users from March to June by underlying cause of death and year

**Figure 3b:**
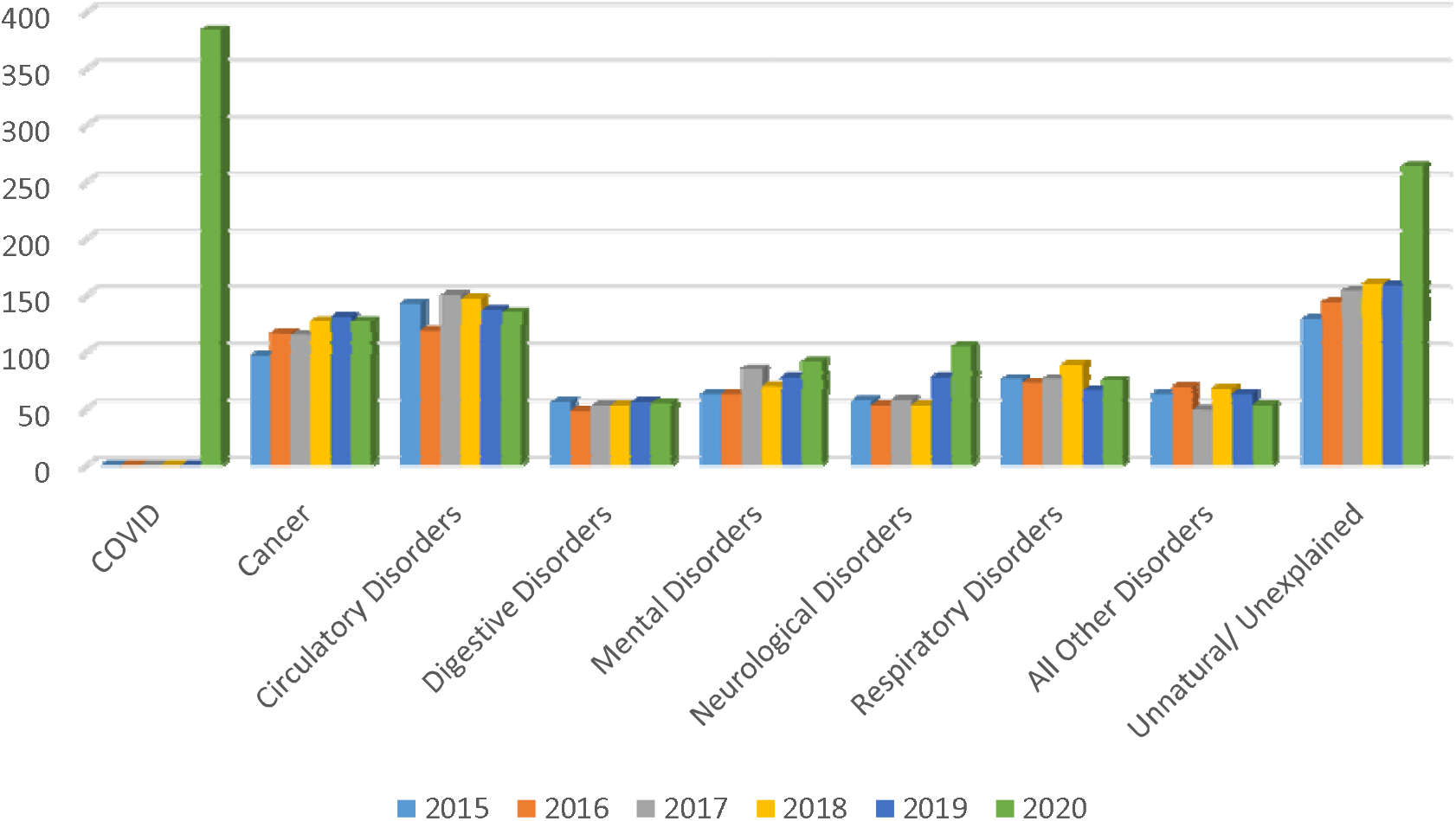
Numbers of deaths in male SLaM service users from March to June by underlying cause of death and year

Considering recency of contact with SLaM (Figures 4a and 4b), most COVID-19 deaths (N=590, 83.3%) were in past rather than current service users. The excess mortality from mental/neurological causes in 2020 was more evident in past service users, while the excess from unnatural/unexplained causes was more evident in current service users. Considering further the excess mortality in current service users, there were 502 deaths in March-June 2020. This number was 111 higher than the average deaths in previous years (391), and this excess was more than accounted for by the 118 deaths from COVID-19 in current service users and the 116 excess deaths from unnatural/unexplained causes (187 in 2020 compared to an average of 71 for 2015-19). For discharged SLaM service users, there were 1001 excess deaths in 2020 (2062 deaths compared to an average of 1061 for 2015-19), with the COVID-caused deaths (n=590) potentially accounting for 59.0% and the 112 excess unnatural/unexplained deaths (n=295 in 2020 compared to the average of 183 in 2015-19) accounting for 11.2%.

**Figure 4a:**
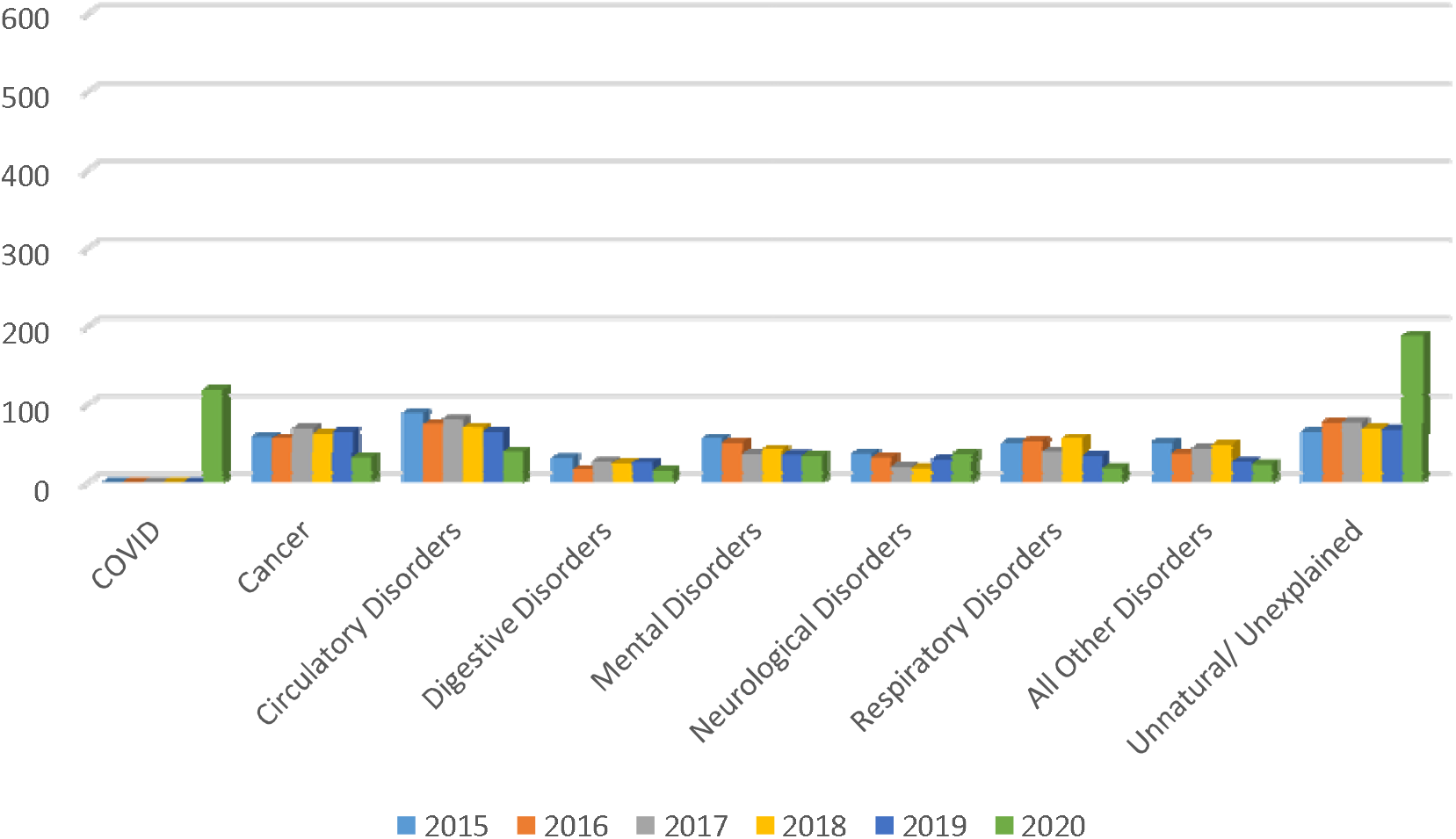
Numbers of deaths in active SLaM service users from March to June by underlying cause of death and year

**Figure 4b:**
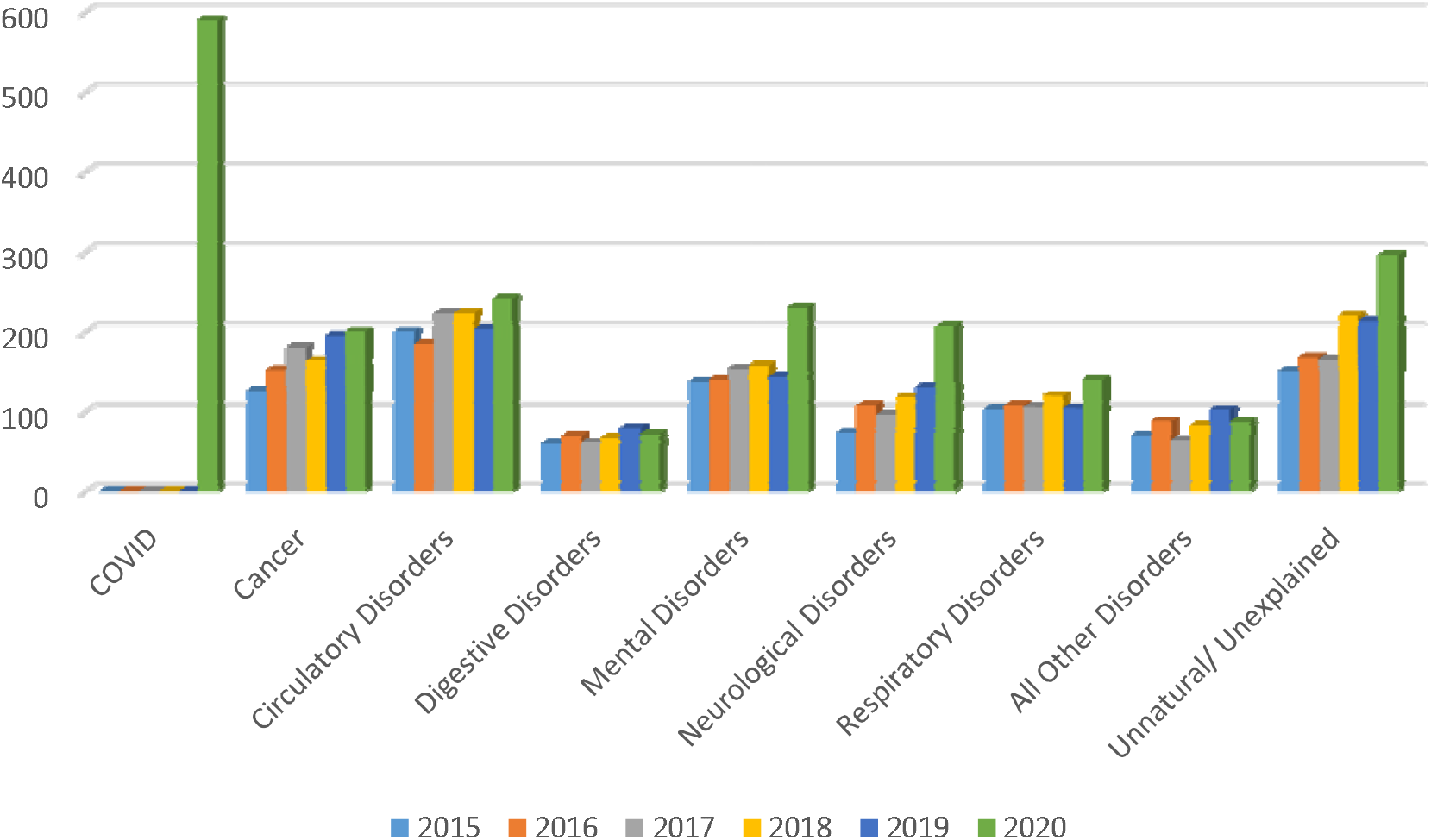
Numbers of deaths in discharged SLaM service users from March-June by underlying cause of death and year

Stratification by ethnic group (Figures 4a-c, Figures 5a-c) showed a higher proportion of deaths that were caused by COVID-19 in patients of Black African/Caribbean ethnicity (37% of deaths, compared to 26% in White British and 27% in those from other ethnic groups), and a higher proportion of unnatural/unexplained deaths in those from other ethnic groups (24%, compared to 17% in White British and 16% in Black African/Caribbean patients).

**Figure 5a:**
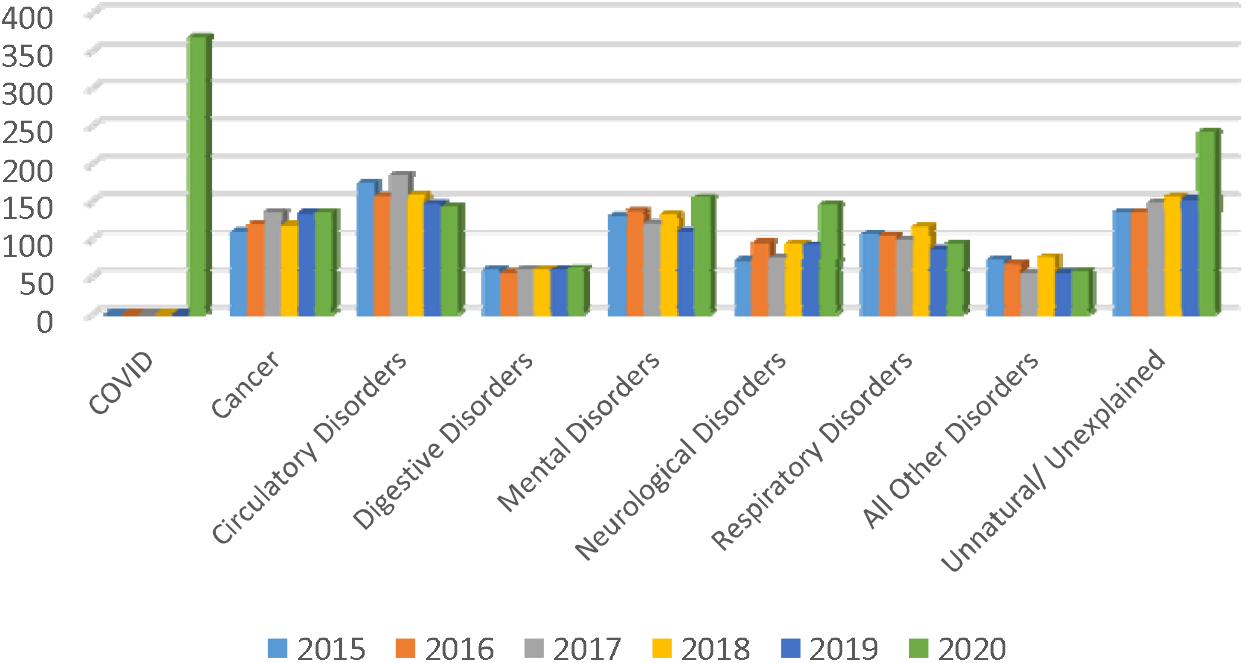
Cause specific mortality -White British

**Figure 5b:**
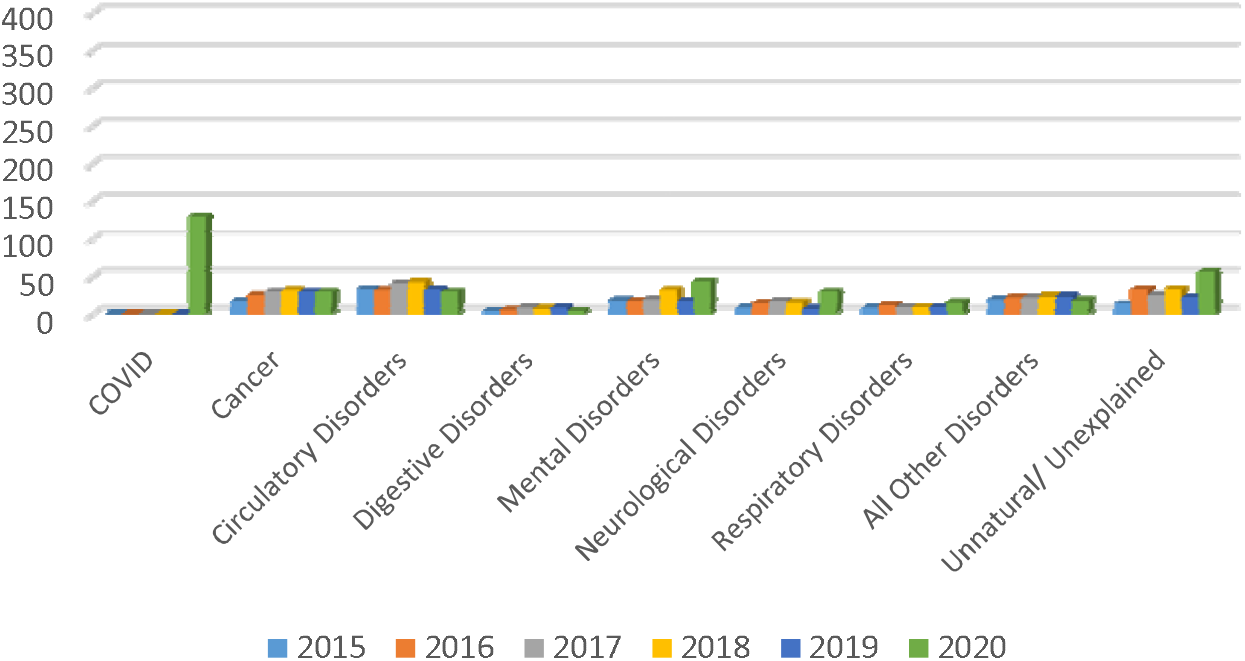
Cause specific mortality - Black African/Caribbean

**Figure 5c:**
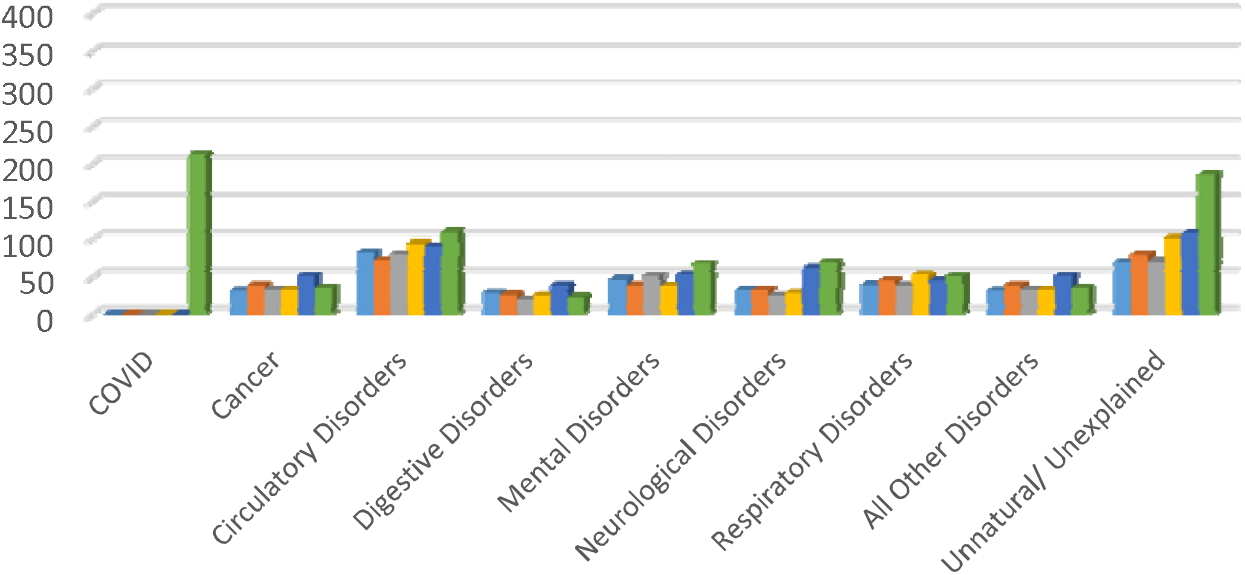
Cause specific mortality - Other ethnic group

**FIGURE 5A.**
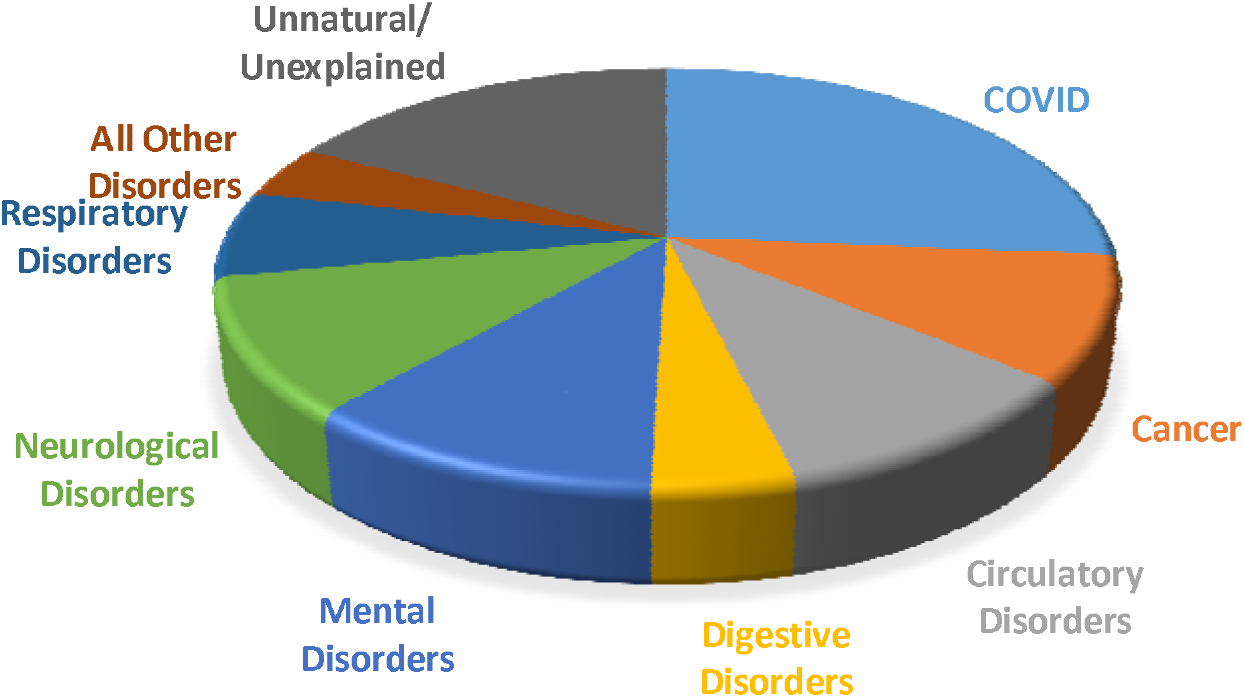
CAUSES OF DEATH -WHITE BRITISH

**FIGURE 5B.**
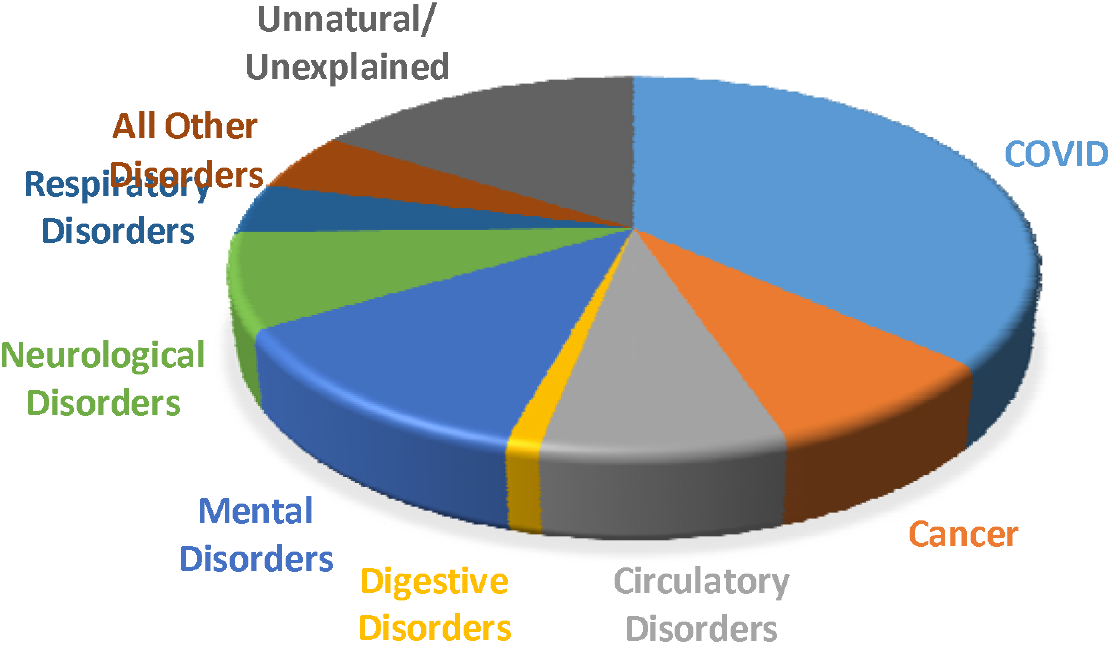
CAUSE OF DEATH – BLACK AFRICAN/CARIBBEAN

**FIGURE 5C:**
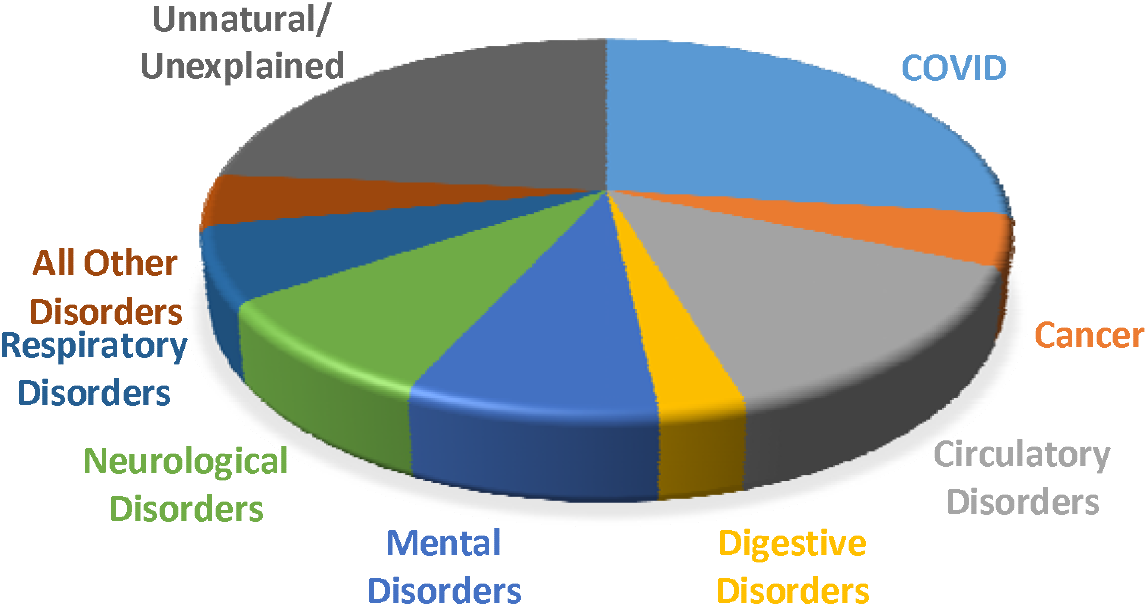
CAUSE OF DEATH - OTHER ETHNIC GROUP

A further description of deaths with mental/neurological disorder as an underlying cause is displayed in Table 1, illustrating that the vast majority in all years (and the excess in 2020) were in the F0x (organic disorders) category for mental disorders and the G3x (other degenerative diseases) category for neurological disorders. In a further analysis of 458 deaths in 2020 with F0x or G3x as an underlying cause of death, mentions of COVID anywhere in the death certificate were only found in 20 (4.4%).

**Table 1:**
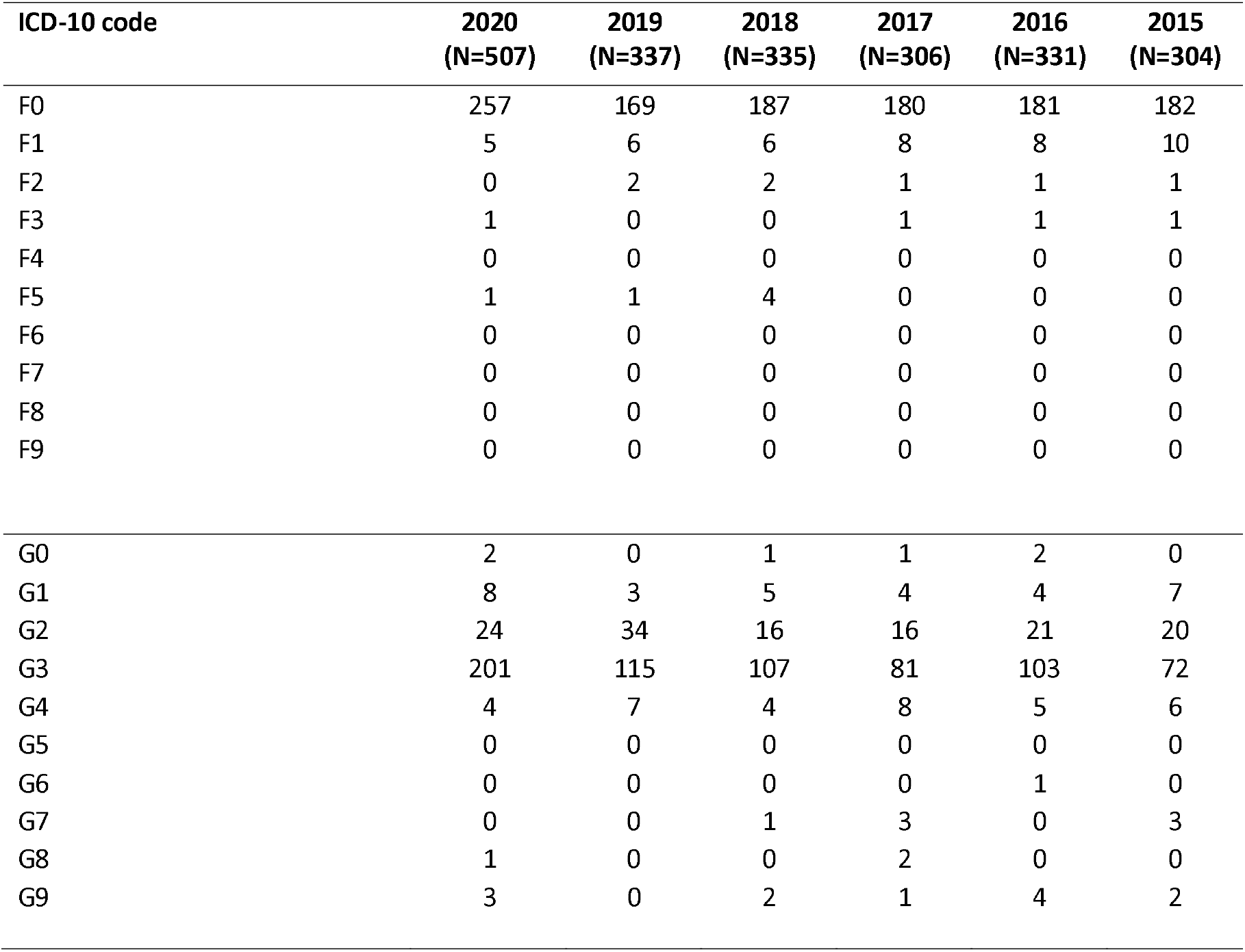
Distribution of deaths with mental disorder (ICD-10 F) or neurological disorder (ICD-10 G) as an underlying cause by next numeric ICD-10 digit

Considering deaths in 2020 in the unnatural/unknown category, numbers ‘unknown’ (i.e. without a registered cause) are displayed by week in Figure 6, showing an increase towards the end of the evaluated period, as well as an excess coinciding with the early weeks of the pandemic period from mid-March to late April. A further breakdown by year is provided in Table 2. Of deaths in 2020, 439 (91.6%) of 479 were in the ‘unknown’ category (i.e. did not have death certification information). Having excluded the 295 deaths (67.2%) where certification would not be expected, the remainder had relatively high proportions awaiting a formal death notice (54.9%) or awaiting an ONS record (13.6%). The 162 awaiting a formal death notice in 2020 represented an excess of 129 compared to the average number for previous years (33): i.e. 11.6% of the 1109 excess total deaths for 2020.

**Table 2:**
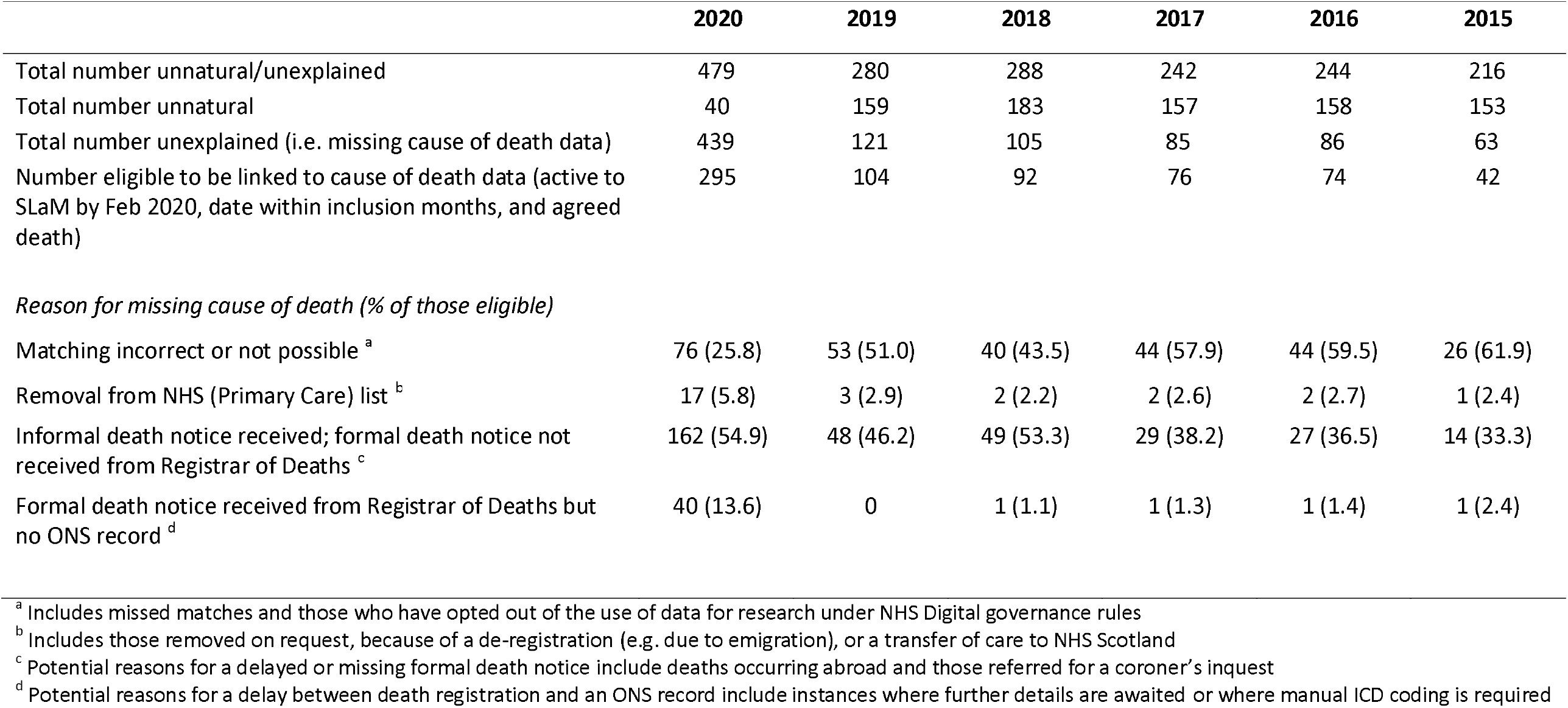
Further description of reasons for deaths classified as unnatural or unexplained.

|  | 2020 | 2019 | 2018 | 2017 | 2016 | 2015 |
| --- | --- | --- | --- | --- | --- | --- |
| Total number unnatural/unexplained | 479 | 280 | 288 | 242 | 244 | 216 |
| Total number unnatural | 40 | 159 | 183 | 157 | 158 | 153 |
| Total number unexplained (i.e. missing cause of death data) | 439 | 121 | 105 | 85 | 86 | 63 |
| Number eligible to be linked to cause of death data (active to SLaM by Feb 2020, date within inclusion months, and agreed death) | 295 | 104 | 92 | 76 | 74 | 42 |
| *Reason for missing cause of death (% of those eligible)* |  |  |  |  |  |  |
| Matching incorrect or not possible <sup>a</sup> | 76 (25.8) | 53 (51.0) | 40 (43.5) | 44 (57.9) | 44 (59.5) | 26 (61.9) |
| Removal from NHS (Primary Care) list <sup>b</sup> | 17 (5.8) | 3 (2.9) | 2 (2.2) | 2 (2.6) | 2 (2.7) | 1 (2.4) |
| Informal death notice received; formal death notice not received from Registrar of Deaths <sup>c</sup> | 162 (54.9) | 48 (46.2) | 49 (53.3) | 29 (38.2) | 27 (36.5) | 14 (33.3) |
| Formal death notice received from Registrar of Deaths but no ONS record <sup>d</sup> | 40 (13.6) | 0 | 1 (1.1) | 1 (1.3) | 1 (1.4) | 1 (2.4) |

<sup>a</sup> Includes missed matches and those who have opted out of the use of data for research under NHS Digital governance rules
<sup>b</sup> Includes those removed on request, because of a de-registration (e.g. due to emigration), or a transfer of care to NHS Scotland
<sup>c</sup> Potential reasons for a delayed or missing formal death notice include deaths occurring abroad and those referred for a coroner's inquest
<sup>d</sup> Potential reasons for a delay between death registration and an ONS record include instances where further details are awaited or where manual ICD coding is required

**Figure 6:**
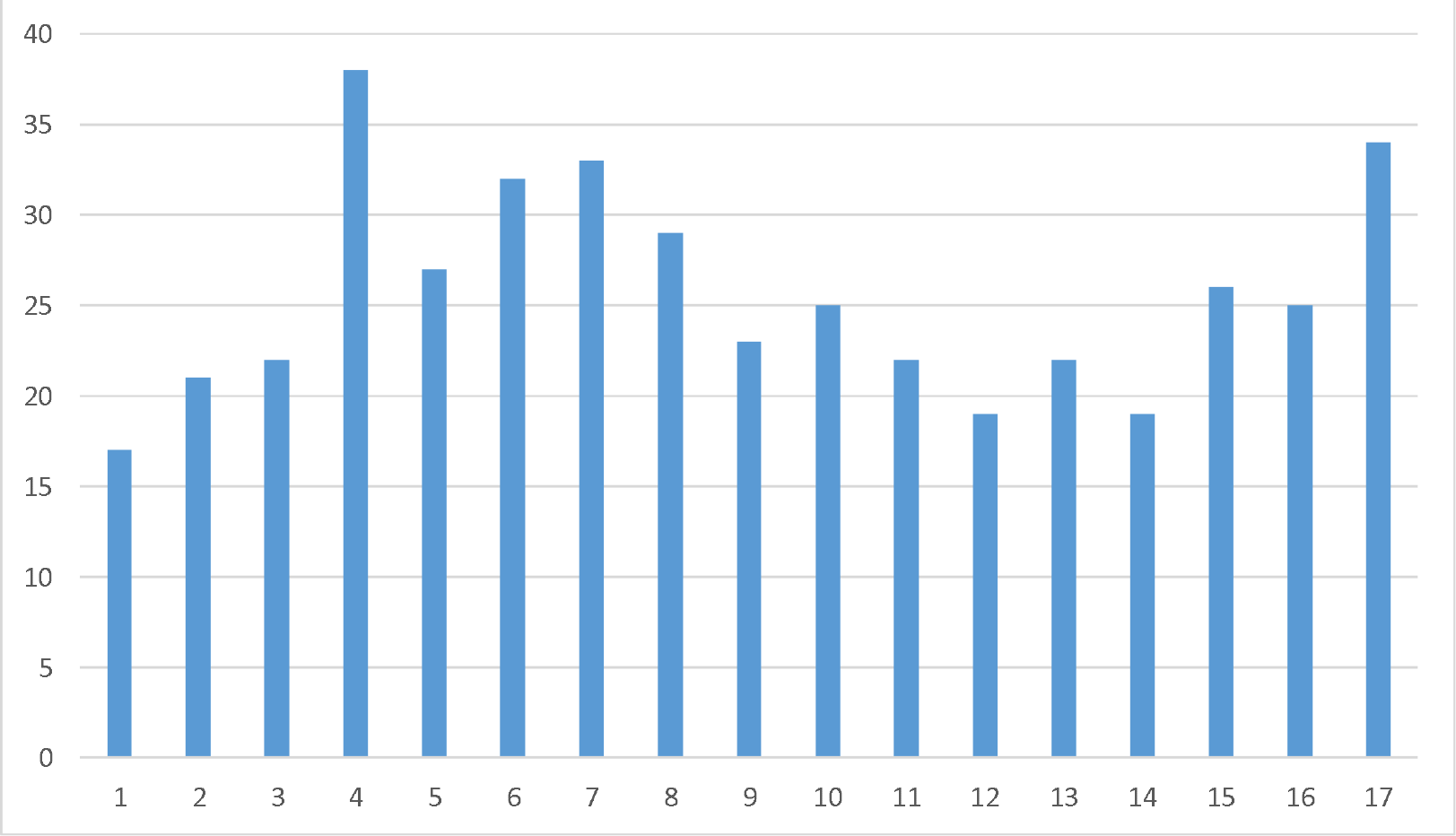
Number of deaths without a registered cause by week from March to June 2020. Note: Week 4 commences 22^nd^ March; Week 12 commences 18^th^ May

## Discussion

We present descriptions of recorded underlying cause of death in a population of past and current mental health service users for the March-June period in 2020, coinciding with the peak of the first COVID-19 wave in London, and compare these with the same period in the five preceding years. Aside from deaths recorded as caused by COVID-19, excess mortality in 2020 was accounted for by unnatural/unexplained causes, and those recorded as caused by a mental/neurological disorder. No excess was seen in any of the other categories.

Deaths with COVID-19 as an underlying cause accounted for close to two thirds of the excess mortality seen in 2020 compared to 2015-19. The majority (around three quarters) of these deaths occurred in service users aged 70+, and over 80% occurred in previous rather than current service users; however, COVID-19 deaths accounted for a substantially higher proportion of excess deaths in current compared to previous service users.

Mental/neurological disorders recorded as underlying causes of death were almost exclusively in organic/neurodegenerative subcategories respectively and the 2020 excess in these causes was exclusively in service users aged 70+ and in those who were discharged rather than receiving current care in March 2020; they were also more evident in female compared to male service users. There were very few with COVID-19 recorded anywhere else on the death certificate.

The broad category of unnatural and unexplained deaths showed an excess overall in 2020, with fewer deaths specifically allocated to ‘unnatural’ categories, but larger numbers with missing cause of death. Of those deaths without a recorded cause, the majority had not yet received a formal death notice – a category which includes those deaths awaiting a coroner’s inquest. Understandably, missing causes of death were increased in the weeks closer to the extraction date; however, notably they were also more common around the early period of the pandemic in London.

Considering limitations, it is important to bear in mind that the data are derived from a single site. Because complete data are being provided for that site with no hypothetical source population intended, calculation of confidence intervals was not felt to be appropriate for the descriptive data provided in this report; applicability to other mental healthcare providers cannot therefore be inferred and would need specific investigation. We additionally aim to provide further analysed output on mortality trends and predictors in future submissions for peer-reviewed publications. Data stratifications were relatively broad, pragmatically driven by the need for sufficiently large group sizes. For example, the conflation of ethnic groups into the categories defined here is inevitably approximate and may fail to reflect varying experiences between constituent groups. Finally, the way in which ‘active’ service users were defined for stratification might have weighted the profile of deaths in this group towards those occurring later in each March-June period, of potential relevance to 2020 mortality if this altered across the first wave of the pandemic.

## Supporting information

STROBE checklist

## Data Availability

Data are available on request from the corresponding author.

## References

1. Chang CK, Hayes RD, Perera G, Broadbent MTM, Fernandes AC, Lee WE, et al. Life expectancy at birth for people with serious mental illness, substance use disorders, and depressive disorders from a secondary mental health care case register in London, UK. PLoS One. 2011; 6: e19590.

2. Woodhead C, Ashworth M, Broadbent M, Callard F, Hotopf M, Schofield P, et al. Cardiovascular disease treatment among severe mental illness patients: a data linkage study between primary and secondary care. British Journal of General Practice. 2016; 66: e374–e81.

3. Woodhead C, Cunningham R, Ashworth M, Barley E, Stewart R, Henderson M. Cervical and breast cancer screening uptake among women with serious mental illness: a data linkage study. BMC Cancer. 2016; 16: 819.

4. Holmes EA, O’Connor RC, Perry VH, Tracey I, Wessely S, Arseneault L, et al. Multidisciplinary research priorities for the COVID-19 pandemic: a call for action for mental health science. Lancet Psychiatry. 2020; Apr 15: https://doi.org/10.1016/S2215-0366(20)30168-1.

5. Hotopf M, Bullmore E, O’Connor RC, Holmes EA. The scope of mental health research during the COVID-19 pandemic and its aftermath.. British Journal of Psychiatry. 2020; 217: 540–2.

6. Stewart R, Martin E, Broadbent M. Mental health service activity during COVID-19 lockdown: South London and Maudsley data on working age community and home treatment team services and mortality from February to mid-May 2020. medRxiv. 2020; 16 Jun 2020: https://www.medrxiv.org/content/10.1101/2020.06.13.20130419v1

7. Stewart R, Broadbent M, Das-Munshi J. Excess mortality in mental health service users during the COVID-19 pandemic described by ethnic group: South London and Maudsley data. medRxiv. 2020; 14 Jul 2020.

8. Stewart R, Soremekun M, Perera G, Broadbent M, Callard F, Denis M, et al. The South London and Maudsley NHS Foundation Trust Biomedical Research Centre (SLAM BRC) Case Register: development and descriptive data. BMC Psychiatry. 2009; 9: 51.

9. Perera G, Broadbent M, Callard F, Chang CK, Downs J, Dutta R, et al. Cohort profile of the South London and Maudsley NHS Foundation Trust Biomedical Research Centre (SLaM BRC) Case Register: current status and recent enhancement of an Electronic Mental Health Record derived data resource. BMJ Open. 2016; 6: e008721.

10. Fernandes AC, Cloete D, Broadbent MTM, Hayes RD, Chang CK, Roberts A, et al. Development and evaluation of a de-identification procedure for a case register sourced from mental health electronic records. BMC Medical Informatics and Decision Making. 2013; 13: 71.

